# Combining clinical and polygenic risk improves stroke prediction among individuals with atrial fibrillation

**DOI:** 10.1101/2020.06.17.20134163

**Authors:** Jack W. O’Sullivan, Anna Shcherbina, Johanne M Justesen, Mintu Turakhia, Marco Perez, Hannah Wand, Catherine Tcheandjieu, Shoa L. Clarke, Robert A. Harrington, Manuel A. Rivas, Euan A Ashley

## Abstract

**Background:** Atrial fibrillation (AF) is associated with a five-fold increased risk of ischemic stroke. A portion of this risk is heritable, however current risk stratification tools (CHA2DS2-VASc) don’t include family history or genetic risk. We hypothesized that we could improve ischemic stroke prediction in patients with AF by incorporating polygenic risk scores (PRS).

**Objectives:** To construct and test a PRS to predict ischemic stroke in patients with AF, both independently and integrated with clinical risk factors.

**Methods:** Using data from the largest available GWAS in Europeans, we combined over half a million genetic variants to construct a PRS to predict ischemic stroke in patients with AF. We externally validated this PRS in independent data from the UK Biobank (UK Biobank), both independently and integrated with clinical risk factors.

**Results:** The integrated PRS and clinical risk factors risk tool had the greatest predictive ability. Compared with the currently recommended risk tool (CHA2DS2-VASc), the integrated tool significantly improved net reclassification (NRI: 2.3% (95%CI: 1.3% to 3.0%)), and fit (χ2 P =0.002). Using this improved tool, >115,000 people with AF would have improved risk classification in the US. Independently, PRS was a significant predictor of ischemic stroke in patients with AF prospectively (Hazard Ratio: 1.13 per 1 SD (95%CI: 1.06 to 1.23))). Lastly, polygenic risk scores were uncorrelated with clinical risk factors (Pearson’s correlation coefficient: −0.018).

**Conclusions:** In patients with AF, there appears to be a significant association between PRS and risk of ischemic stroke. The greatest predictive ability was found with the integration of PRS and clinical risk factors, however the prediction of stroke remains challenging.

## Introduction

Atrial fibrillation (AF) is the most common cardiac arrhythmia and its prevalence is increasing (1). Atrial fibrillation itself can cause substantial morbidity, including a 5-fold increased risk of ischemic stroke (2).

To help prevent the thromboembolic complications of AF, selected patients are offered prophylactic anticoagulation. This prophylaxis is highly effective in the right patient (3–5), but the selection of these patients remains difficult (6, 7). The current gold standard risk stratification tool is an amalgamation of clinical risk factors (CHA2DS2-VASc) (4). However, there are limitations in the development, validation and performance of CHA2DS2-VASc. Most notably the small number of AF patients in the development (n=1,084) (6), and short follow up, small numbers, and conflicting performance in validation studies (8). Additionally, CHA2DS2-VASc tool does not include family history or genetic risk of ischemic stroke, despite evidence suggesting the risk of ischemic stroke is heritable (∼40% heritability) (9). Previous research has shown that polygenic risk scores are comparable to clinical risk factors in the prediction of ischemic stroke in the general population (10), however this has not been extended into patients with AF, nor did it examine CHA2DS2-VASc.

Given the known heritability of ischemic stroke, and the apparent need to improve the existing gold standard risk tool (CHA2DS2-VASc), we set out to construct a polygenic risk score (PRS), and then an integrated genetic and clinical risk tool (CHA2DS2-VASc + PRS) to help predict which patients with AF will go on to develop ischemic stroke.

## Methods

### Study design

We followed a similar study design to previously published PRS papers (10–14); in line with recommended methodological (15) and reporting guidance (16). We will briefly describe the five broad steps we completed in this paragraph (Figure1), and then we elaborate on each of these steps individually in the below paragraphs. The five steps were: 1. Curation of previously published GWAS summary statistics, 2. Accounting for linkage disequilibrium (LD) in GWAS summary statistics, using the R package *lassosum* (17) 3. Construction of PRS (see eMethods) in our UK Biobank *prevalent* cohort. Eighty different PRS were constructed across the lassosum hyperparameters (λ and s). 4. Determining the most accurate PRS in the UK Biobank *prevalent* Cohort. 5. The PRS with the greatest predictive accuracy (from step 4) was then validated in the UK Biobank *incident* cohort.

**Figure 1:**
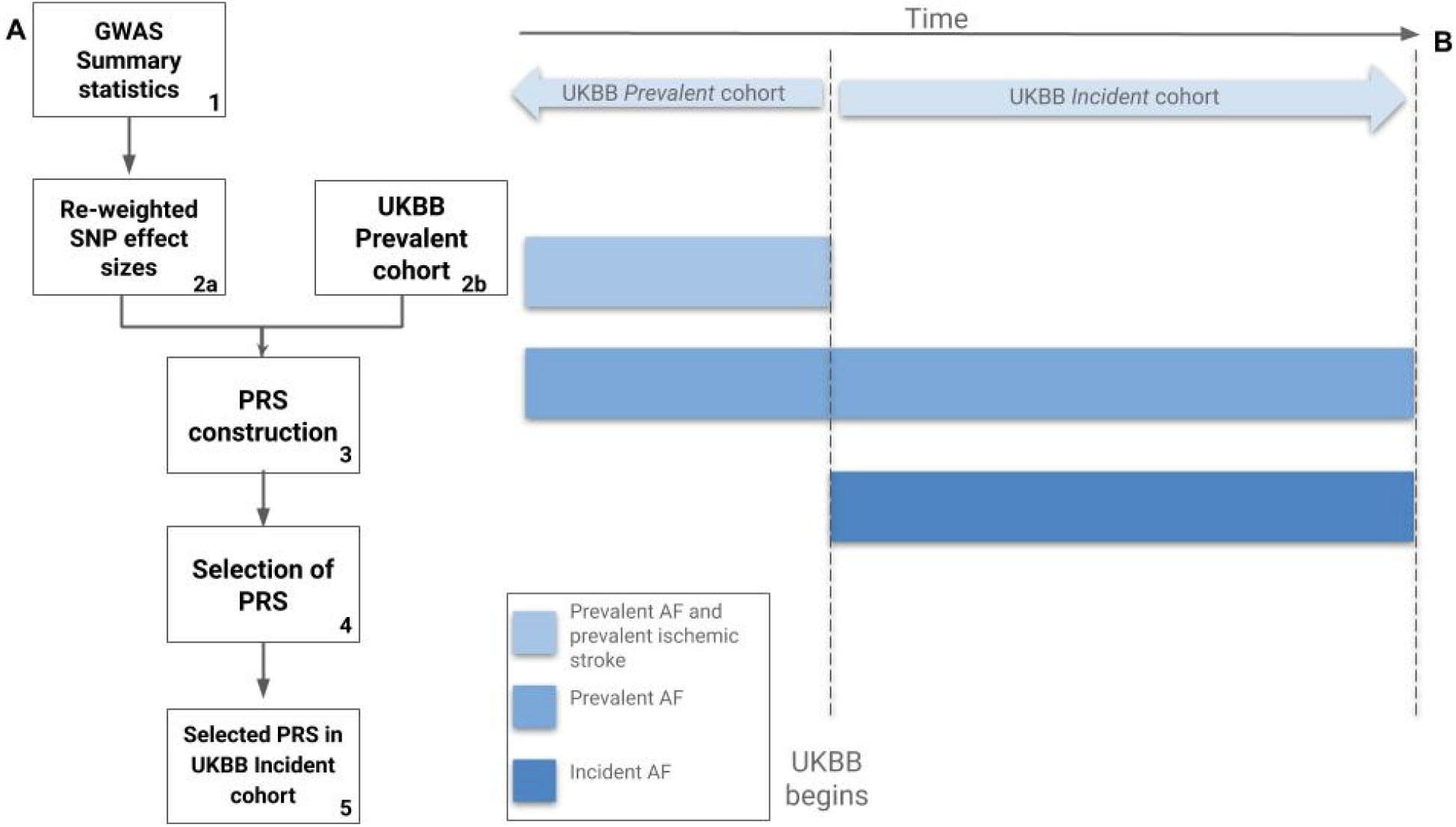
A represents the five broad study steps. B displays the characteristics of the participants in the UK Biobank *prevalent* Cohort and the UK Biobank *incident* Cohort, in relation to their AF diagnosis.

We attained GWAS summary statistics from the MEGASTROKE consortium (http://www.megastroke.org/). This GWAS was performed on 446,696 participants (40,585 cases (stroke); 406,111 noncases (no stroke)) and stratified results by ancestry and stroke sub-type (18). We used the GWAS summary statistics corresponding to cardioembolic stroke for participants of European ancestry (18).

To account for LD, we used *lassosum* to re-weight SNV effect sizes from the MEGASTROKE GWAS summary statistics (17). The importance of accounting for LD has been well-described previously (11, 12, 19), as has the lassosum algorithm to account for LD (12, 17). Briefly, lassosum is a machine learning approach that reweights SNV effect sizes via a penalized regression model, which essentially shrinks a proportion of the SNV effect-size to zero. A reference population is used, in our case we used the LD regions defined in Berisa et al (20) The model is tuned via model tuning parameters (λ and s - penalty terms in the model which define how many SNVs are set to zero), and 80 versions of re-weighted SNVs are created across these tuning parameters (there are 80 combinations of tuning parameters). Polygenic risk scores are then constructed using these re-weighted SNVs by summing the SNV weights per allele (0, 1 or 2) per participant (see eMethods). As there are 80 variations of re-weighted SNVs across the tuning parameters, 80 polygenic risk scores are produced per participant. These 80 scores were then trained in our UK Biobank *prevalent* cohort. Our UK Biobank *prevalent* cohort consisted of participants with a history of AF followed by a history of ischemic stroke before the beginning of the UK Biobank, and an identical number of randomly selected AF controls: participants with AF, but who did not suffer from an ischemic stroke before the beginning of the UK Biobank. The PRS score with the best performance was selected (17). We chose to use lassosum as it has been shown to have greater predictive performance over other methods, including LDpred (12, 17, 21).

Once the best performing PRS is selected, we tested the predictive ability of this PRS (along with other covariates) in the entirely independent UK Biobank *incident* cohort. The *incident* cohort consists of UK Biobank participants with either a) a history of AF before the UK Biobank begun (prevalent AF), but who had not suffered an ischemic stroke at UK Biobank recruitment, or b) a history of AF after they were enrolled in the UK Biobank (incident AF), who had also not had an ischemic stroke between UK Biobank enrolment and the development of AF. There was no overlap of participants between the UK Biobank *prevalent* and *incident* cohort. Similarly, The MEGASTROKE GWAS was entirely independent from the UK Biobank *incident* and *prevalent* cohorts.

In the same cohort (UK Biobank *incident* cohort), we examined the predictive ability of clinical risk factors for ischemic stroke in patients with AF, both individually and collectively (via the commonly used tool: CHA2DS2-VASc). Lastly, we examined a risk tool that integrates clinical risk factors with PRS (we refer to this as CHA2DS2-VASc -G).

### Study population

All participants included in our study were from the UK Biobank (UK Biobank). We included UK Biobank participants of European descent with a history of atrial fibrillation (AF), either before enrollment in the UK Biobank (prevalent AF) or after enrollment in the UK Biobank (incident AF). All UK Biobank participants were recruited between 2006 and 2010, and (at the time of recruitment) were aged 40 to 69. Recruited participants were assessed at dedicated assessment centers across the UK. At assessment centers, historical medical records were collected (self-reported and via electronic health record), and participants were prospectively followed (22). The UK Biobank received ethical approval from the National Health Service’s National Research Ethics Service North West (11/NW/0382).

After attainment of all participants with AF, participants were excluded if they had missing genetic data (see *Genetic Data* below), were not of European descent, withdrew consent, or had suffered an ischemic stroke before the development of AF. The remaining participants were divided into two cohorts; the UK Biobank *prevalent* cohort and the UK Biobank *incident* cohort. As described above, the UK Biobank *prevalent* cohort (n=566) consists of participants with a history of prevalent AF (before UK Biobank) and a history of ischemic stroke before the beginning of the UK Biobank, as well as an identical number of randomly selected controls (participants with AF, without an ischemic stroke before the beginning of the UK Biobank). We used this cohort to tune and then select the most accurate PRS of all those created with lassosum - this ensures that all of participants with incident ischemic stroke are in the *incident* cohort, maximising power for Cox regression (a similar method to Elliot et al (12)). The selected PRS was then validated in the UK Biobank *incident* cohort.

The UK Biobank *incident* cohort contains the remaining patients with AF (n=15,412). In this independent cohort we assessed the predictive accuracy of the PRS, as well as the clinical risk factors (CHA2DS2-VASc) and the integrated clinical and genomic risk tool (CHA2DS2-VASc - G). We assessed the risk of ischemic stroke in patients with an established diagnosis of AF. Thus, for both our *prevalent* and *incident cohorts* participants with AF who had suffered an ischemic stroke at the end of follow up constituted ‘cases’ and participants with AF who had *not* suffered an ischemic stroke at the end of follow up constituted ‘noncases’ (i.e. all included participants had AF, the outcome of interest was ischemic stroke in patients with AF). The codes used to define AF, as well as ischemic stroke are defined in the appendix (see appendix, eTable1).

### Genetic data

The UK Biobank genotyping and imputation techniques have been extensively described previously (23, 24). In summary, initially custom Affymetrix arrays were used to genotype participants for the UK Biobank Lung Exome Variant Evaluation study, and subsequently the UK Biobank Axiom array was used (23, 24). Imputation was computed centrally using IMPUTE2 (25) based on a combined sample of UK10K sequencing and 1000 Genomes Project imputation reference panels. Genetic principal components analysis (PCA) was also performed by the UK Biobank.

We performed quality control on both the UK Biobank data and the MEGASTROKE GWAS summary statistics in line with recommended guidance (15). On the UK Biobank data, we included SNVs that met the following quality control filters: minor allele frequency (MAF) > 0.001, P-value from Hardy-Weinberg Equilibrium Fisher’s exact or chi-squared test > 1e-6, SNVs that were present in >99% of included participants (Europeans), an INFO score >99, non-duplicate, and non-mismatching. We also excluded individuals with > 1% genotype missingness, kinship cutoff > 0.125, and heterozygosity 3 standard deviations from the mean. Further, duplicate and mismatching SNVs were removed from the MEGASTROKE GWAS. Genetic filtering analyses were conducted in PLINK (version 2), Python (version 3.7.3), and R (version 3.6.2).

### Clinical risk score

To compare our constructed PRS with the clinical risk factors used to predict ischemic stroke in those with AF, we identified if included participants had been diagnosed with (at or before their diagnosis of AF): heart failure, hypertension, vascular disease (coronary artery disease, peripheral vascular disease, and or atherosclerosis), thromboembolism and/or diabetes (type 1 or 2). We also defined the participant’s age (at AF diagnosis), sex assigned at birth, and self-reported history of having been prescribed warfarin. These clinical risk factors were selected to enable us to construct the currently recommended clinical tool to assess the risk of ischemic stroke in patients with AF: CHA2DS2-VASc (4). The codes used to define these clinical risk factors are listed in the appendix (eTable1). We also calculated each participant’s CHA2DS2-VASc score at AF diagnosis, and defined a variable to indicate whether prophylactic anticoagulation should be recommended in line with guidelines (e.g. ‘yes’ if CHA2DS2-VASc score of ≥2 in men and ≥3 in women) (4).

### Statistical Analysis

We completed the following broad steps: First, we investigated the predictive ability of PRS in logistic regression models. Second, we assessed the correlation between PRS and CHA2DS2-VASc scores, and third we investigated the ability of PRS to predict incident ischemic strokes via Cox-regression models (starting follow up from AF diagnosis). The PRS was scaled to a mean of 0 and a standard deviation of 1 to facilitate interpretation, in line with current literature. All reported analyses were conducted on the UK Biobank *incident* cohort, which represents our validation cohort - entirely independent from the GWAS summary statistics and the UK Biobank *prevalent* cohort.

We constructed logistic regression models examining PRS, controlling for age at recruitment, sex, UK Biobank array type, and the first 10 principal components (PCs). We report the odds ratio (OR) for PRS, as well as the area under the ROC curve (AUROC) for the model. We conducted further sensitivity analyses: logistic regression models including the aforementioned covariates as well as 1. The presence of warfarin prescription (additional covariate), 2. Excluding participants who had been prescribed warfarin, and 3. CHA2DS2-VASc score (additional covariate). We conducted sensitivity analyses controlling for warfarin as it is often prescribed to patients with AF and may mitigate their risk of ischemic stroke. We also constructed a logistic regression with CHA2DS2-VASc score as the sole predictor, to enable comparison between the model with both PRS and CHA2DS2-VASc.

We then assessed the correlation between the participant’s CHA2DS2-VASc score (at AF diagnosis) and PRS using Pearson’s Correlation Coefficient (and as sensitivity analyses, Spearman’s and Kendall’s tau (rank)).

To predict incident ischemic strokes over time, we constructed Cox-regression models adjusting for sex, array and the first 10 PCs (we set out to adjust for age, but its inclusion violated the hazard assumption - we conducted sensitivity analyses using Age as time scale to address this, see below and appendix). We constructed the following models: 1. Using PRS as a sole predictor. 2. Adjusting for warfarin prescription, 3. Excluding those prescribed warfarin, 4. PRS and CHA2DS2-VASc score, 5. CHA2DS2-VASc score as the sole predictor, and 6. Individual components of CHA2DS2-VASc score as predictors (e.g. Heart failure diagnosis). For all models the proportionality assumption was assessed via the global test for scaled Schoenfeld residuals (no models violated the global assumption unless otherwise stated) and calibration was assessed via the Greenwood-Nam-D’Agostino χ2 test (26). We also conducted sensitivity analyses using age as the time scale (sensitivity analysis 1), which did not violate the hazard assumption. We report a variety of model fit and accuracy metrics: likelihood ratio χ2 test, LJ-index, and Net Reclassification Improvement (NRI). We calculated NRI using a risk threshold cut-off of 4% (to define high and low risk threshold, reflecting those eligible for anticoagulation (high) and those not eligible (low)). The NRI metric (27), is the sum of the net reclassification improvement in cases (the proportion of cases correctly reclassified as high risk minus those incorrectly reclassified as low risk) and the net reclassification improvement in noncases (the proportion of noncases correctly reclassified as low risk minus those incorrectly reclassified as high risk). We chose the risk threshold cutoff of 4% in line previously literature (28). This threshold cutoff was also supported by our results; we found that the risk of stroke in those eligible for anticoagulation and not eligible (as determined by CHA2DS2-VASc score) reached a threshold of around 4% (see table 2). As a sensitivity analysis we calculated NRI using a 5% risk threshold (Sensitivity analysis 2).

**Table 1:** Descriptive characteristics of UK Biobank *incident* Cohort at AF diagnosis. HTN = Hypertension, AF = Atrial Fibrillation

|  | **All**<br>**(n=15,412)** | **Ischemic**<br>**stroke (n =**<br>**672)** | **No ischemic**<br>**stroke**<br>**(n=14,740)** |
| --- | --- | --- | --- |
| Age at recruitment<br>[mean, (sd)] | 62.8 (5.9) | 64.4 (5.3) | 62.7 (5.9) |
| Age at AF diagnosis<br>[mean, (sd)] | 63.8 (8.7) | 66.5 (7.3) | 63.7 (8.8) |
| Sex (% male) | 10,908 (66.6%) | 581 (66.2%) | 10327 (66.6%) |
| CHA2DS2-VASC = 0 | 4292 (26.2%) | 137 (15.6%) | 4155 (26.8%) |
| CHA2DS2-VASC = 1 | 5617 (34.3%) | 280 (31.9%) | 5337 (34.4%) |
| CHA2DS2-VASC = 2 | 4020 (24.5%) | 253 (28.8%) | 3767 (24.3%) |
| CHA2DS2-VASC = 3 | 1714 (10.5%) | 144 (16.4%) | 1570 (10.1%) |
| CHA2DS2-VASC = 4 | 581 (3.6%) | 49 (5.6%) | 532 (3.4%) |
| CHA2DS2-VASC = 5 | 137 (0.8%) | 13 (1.5%) | 124 (0.8%) |
| CHA2DS2-VASC = 6 | 18 (0.1%) | 2 (0.2%) | 16 (0.1%) |
| CHA2DS2-VASC = 7 | 1 (0.01%) | 0 | 1 (0.006%) |
| HTN, No. | 6987 (42.7%) | 456 (51.9%) | 6531 (42.1%) |
| Vascular disease, No. | 1979 (12.1%) | 145 (16.5%) | 1834 (11.8%) |
| Heart Failure, No. | 1642 (10.0%) | 103 (11.7%) | 1539 (9.9%) |
| Diabetes, No. | 1770 (10.8%) | 129 (14.7%) | 1641 (10.6%) |

**Table 2:** Reclassification table comparing classification using conventional risk prediction tool (CHA2DS2-VASc) compared with our integrated genetic and clinical risk factors tool (CHA2DS2-VASc -G)). Up and down arrows denote up or down-classified participants respectively: Up arrow denotes participants who were moved from below the anticoagulation threshold (with CHA2DS2-VASc) to above the anticoagulation threshold (with CHA2DS2-VASc -G). Down-arrow denotes participants who were moved from above the anticoagulation threshold (with CHA2DS2-VASc) to below the anticoagulation threshold (with CHA2DS2-VASc -G). Horizontal arrows represent participants who stay in the same category for both risk tools. The last column shows the observed number of ischemic strokes in the different reclassification groups over follow up.

| **Category** | **CHA2DS2-VASc (%)** | **CHA2DS2-VASc -G (%)** | **CHA2DS2-VASc -G breakdown (%)** | **n Ischemic stroke over f/u (%)** |
| --- | --- | --- | --- | --- |
| Anticoagulation recommended | 4863 (30.5%) | 5310 (33.3%) | 4083 (77.0%) → | 221 (5.4%) |
|  |  |  | 1227 (23.0%) ↑ | 71 (5.8%) |
| Anticoagulation not recommended | 11066 (69.5%) | 10619 (66.7%) | 9839 (92.7%) → | 362 (3.7%) |
|  |  |  | 780 (7.4%) ↓ | 30 (3.9%) |

Additionally, we constructed genomically-enhanced CHA2DS2-VASc scores (what we termed: CHA2DS2-VASc-G), where we added one point if a participant was in the top 25% PRS risk, maintained the observed CHA2DS2-VASc score if a participant was in the middle 50% of PRS risk, and subtracted a point if the participant was in the bottom 25% PRS risk. We did this as the current clinical risk tools for the prediction of ischemic stroke in patients with AF (CHA2DS2-VASc) differs from many other existing clinical risk tools. The CHA2DS2-VASc uses a numeric threshold based on the number of risk factors an individual has, rather than a risk percentage that is used by, for instance, the American Heart Association (AHA) and American College of Cardiology (ACC) Pooled Cohort Equation (PCE) for Atherosclerotic Cardiovascular Disease (ASCVD), where 7.5% risk threshold is used for the consideration of statin prescription. Thus, to display reclassification tables we calculated genomically-enhanced CHA_2_DS_2_-VASc scores, and assigned each participant’s anticoagulation recommendation in line with current guidance (e.g. anticoagulation recommended if CHA_2_DS_2_-VASc score of ≥2 in men and ≥3 in women (11)). We then compared the number of participants that would be above and below the anticoagulation threshold for the conventional CHA_2_DS_2_-VASc score and for the genomically-enhanced CHA_2_DS_2_-VASc score (CHA_2_DS_2_-VASc-G). We compared the proportion of strokes observed in participants who were a) Shared high risk: classified as above the anticoagulation risk (e.g. CHA_2_DS_2_-VASc score of ≥2 in men and ≥3 in women) for both CHA_2_DS_2_-VASc and CHA_2_DS_2_-VASc -G and b) Up-classified: classified as above the anticoagulation risk when using CHA_2_DS_2_-VASc -G, but not CHA_2_DS_2_-VASc. We also compared the participants who were c) Down-classified: classified as *below* the anticoagulation risk when using CHA_2_DS_2_-VASc -G, but not CHA_2_DS_2_-VASc, and d) Shared low risk: classified as below the anticoagulation risk for both CHA_2_DS_2_-VASc and CHA_2_DS_2_-VASc -G. We compared proportions using a two proportion z-test to determine if the up-classified group had a similar proportion of strokes to the shared high risk group, which would give us some indication that our up-classified group was at similar stroke risk to the shared high risk group. We performed a similar analysis (and for the same reasons) between the down-classified and shared low risk group. We conducted two further sensitivity analyses: one removing participants prescribed warfarin (sensitivity analysis 3) and one without subtracting a point for participants in the bottom 25% PRS risk (i.e. only add 1 point to a participant’s CHA2DS2-VASc if they were in the *top* 25% PRS risk) (sensitivity analysis 4).

We conducted the two aforementioned approaches to reclassification: 1. Net Reclassification Index using a 4% risk threshold and 2. Re-calculating each participant’s CHA_2_DS_2_-VASc score with the addition of their PRS. We did this as the CHA_2_DS_2_-VASc score is currently recommended to be used in clinical practice by calculating each participant’s risk factors (method 2), however many other risk tools use a percentage risk threshold (method 1), such as the AHA/ACC PCE for Atherosclerotic Cardiovascular Disease (ASCVD).

## Results

The characteristics of the UK Biobank *incident* cohort are reported in Table 1; there were 15,929 participants with AF (eFigure1), of which 684 suffered an ischemic stroke, and 15,245 did not, over follow up. Participants were followed up for a median of 7 years (interquartile range: 5.9). After re-weighting, 530,933 SNVs had a non-zero effect size and were included in our PRS.

The logistic regression analyses showed an AUROC for CHA_2_DS_2_-VASc of: 0.60 (95%CI: 0.58 to 0.62) (appendix, eTable2), with the addition of PRS this rose to 0.61 (95%CI: 0.59 to 0.63) and corresponded to an PRS odds ratio (OR) of 1.14 per SD (95%CI: 1.06 to 1.23). The analysis using PRS as the sole predictor revealed a PRS OR 1.14 per SD (95%CI: 1.06 to 1.23) (figure 2), and an area under the receiver operator curve (AUROC) of 0.60 (95%CI: 0.58 to 0.62). Warfarin was prescribed to 2,326 AF participants at UK Biobank recruitment. The sensitivity analyses both adjusting for warfarin and removing participants prescribed warfarin revealed modestly improved discrimination (appendix, eTable2), albeit with a loss of power. Further, we found that PRS did not correlate with an individual’s CHA_2_DS_2_-VASc score (Pearson’s correlation coefficient = −0.018, appendix, eTable3).

**Figure 2:**
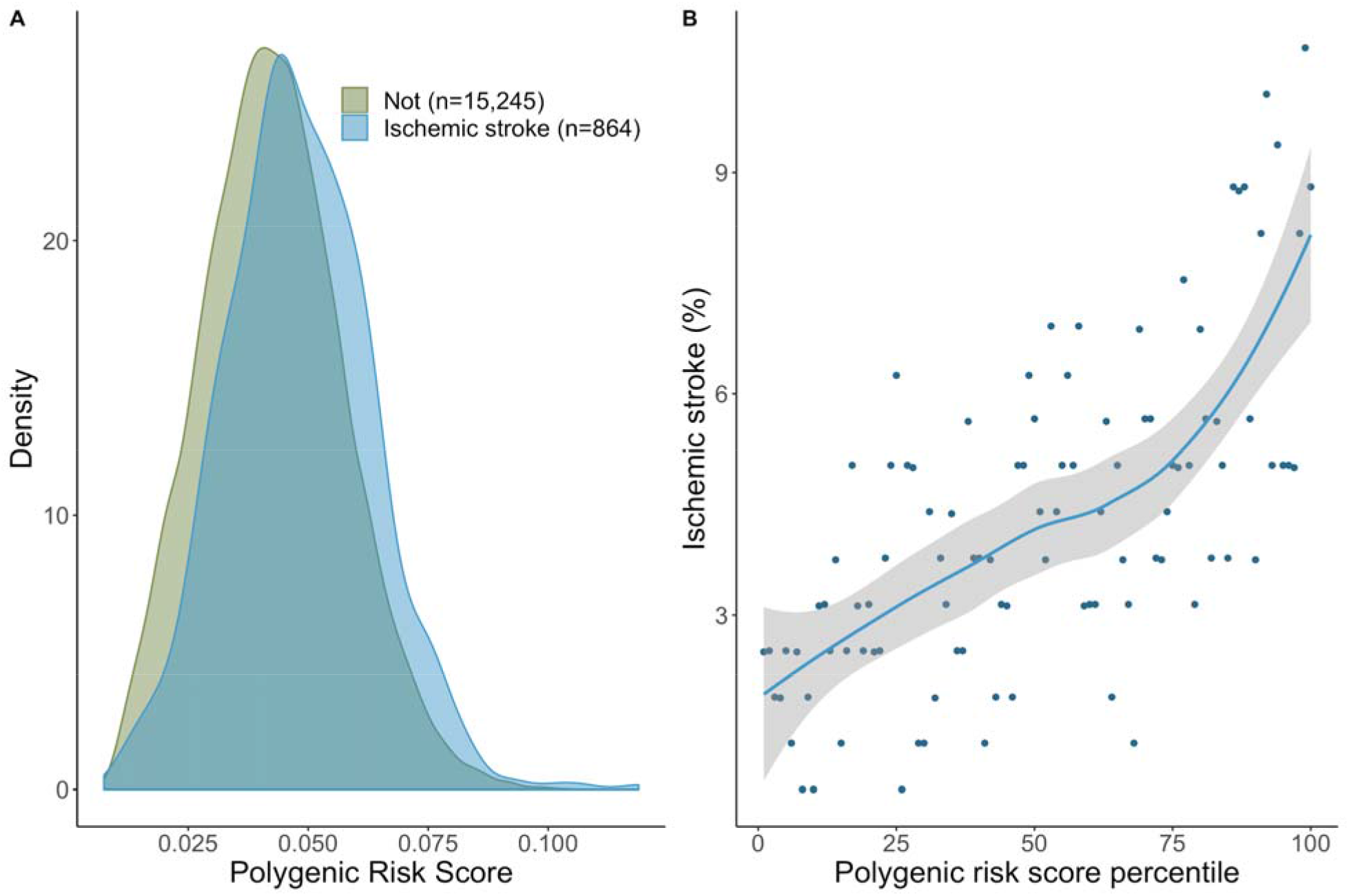
A: Histogram of participants with AF, colour representing those that had an ischemic stroke or not. B: Participants are binned into 100 groups to determine their polygenic risk score (PRS) percentile (x-axis), the prevalence of ischemic stroke (at the end of follow up) is represented on the y axis. For both plots, the PRS is adjusted via logistic regression adjusting for the following covariates: age, sex, first 10 principal component of ancestry, and array platform.

The Cox regression analysis using PRS as the sole predictor revealed a hazard ratio (HR) of 1.13 (95%CI: 1.04 to 1.21) per 1 SD, and a C-statistic of 0.56 (95%CI: 0.54 to 0.58) (figure 3). The same analysis adjusting for age violated the Cox proportional hazards assumption, but nevertheless showed a similar PRS HR (1.14, 95%CI: 1.01 to 1.23), and higher C-statistic (0.63 (95%CI: 0.61 to 0.65). The analysis that adjusted for warfarin prescription violated the Cox proportional hazards assumption, but nevertheless showed a similar HR and C-statistic. The analysis that removed participants prescribed warfarin did not violate the proportional hazard assumption and showed a HR of 1.13 (95%CI: 1.04 to 1.22) and a C-statistic of 0.57 (95%CI: 0.54 to 0.59). The sensitivity analyses using age as time scale for all aforementioned analyses showed largely consistent results (sensitivity analysis 2, eTable 4). All models were well-calibrated (P >0.2 for all models via the Greenwood-Nam-D’Agostino χ2 test).

**Figure 3:**
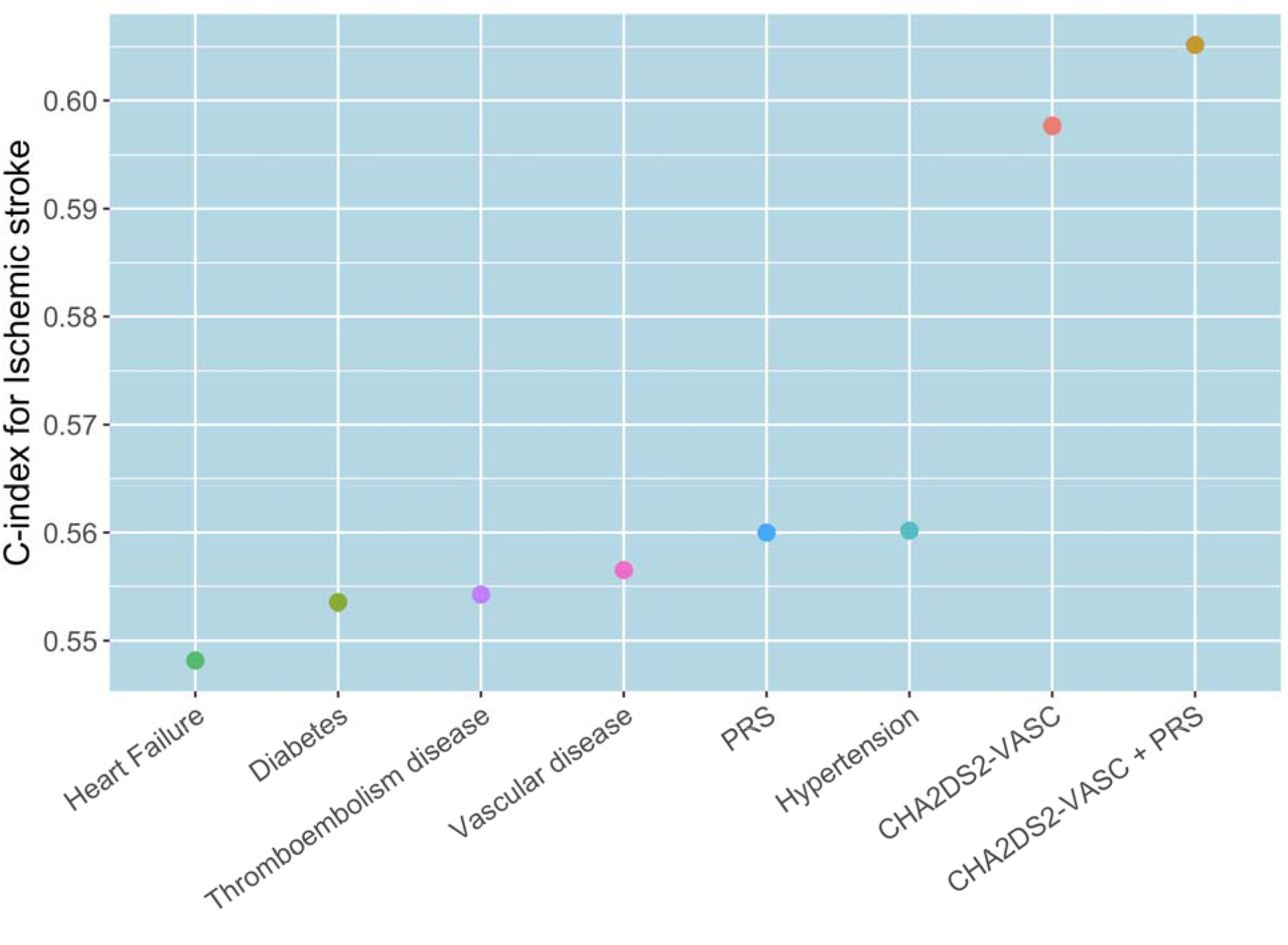
C-statistics for each individual component of CHA2DS2-VASc, as well as for polygenic risk score (PRS), CHA2DS2-VASc collectively and the integrated CHA2DS2-VASc -G (CHA2DS2-VASc and PRS). C-statistics derived from Cox regression models adjusting for sex, array and first 10 Principal Components of ancestry.

The C-statistic for the integrated PRS and CHA_2_DS_2_-VASc model was 0.61 (95%CI: 0.58 to 0.63) (figure 3) and the PRS HR was unchanged (1.13, 95%CI: 1.05 to 1.22). Compared with the currently recommended CHA_2_DS_2_-VASc only model, the integrated PRS and CHA_2_DS_2_-VASc risk model showed a significantly improved statistical fit (χ2 P =0.002), modestly improved discrimination (figure 3) and improved overall Net Reclassification Index (NRI): 2.3% (95%CI: 1.3% to 3.0%) (e Table 5). The NRI (method 1 in *Methods*) was significantly improved for non-cases (2.3% (95%CI: 0.6% to 5.4%) and no different for cases (ischemic strokes) (0.01% (95%CI: −0.4% to 0.1%) (eTable 5). Both models were well calibrated (P >0.3 for both models via the Greenwood-Nam-D’Agostino χ2 test). The sensitivity analysis using a risk threshold of 5% showed a similarly significant overall, but lower NRI (1.91% (0.12% to 6.4%). This NRI was significant for noncases, but not for cases (Sensitivity analysis 2, eTable 6).

For the genomically-enhanced CHA_2_DS_2_-VASc scores (CHA2DS2-VASc-G), the proportion of strokes observed in participants that were up-classified (moved from below the anticoagulation threshold with CHA_2_DS_2_-VASc to above the anticoagulation threshold with CHA_2_DS_2_-VASc - G) was similar (statistically no different) to the proportion of strokes in participants who were classified as high risk in both CHA_2_DS_2_-VASc and CHA_2_DS_2_-VASc -G (above anticoagulation threshold with both): 5.8% vs. 5.4%, P=0.7, table 2, and figure 4 (method 2 in *Methods*). Indicating the up-classified participants had a similar stroke risk to those at shared high risk. Similarly, the proportion of strokes in participants down-classified (moved from above the anticoagulation threshold with CHA_2_DS_2_-VASc to below the anticoagulation threshold with CHA_2_DS_2_-VASc -G) was no different to the proportion of strokes in participants who were classified as low risk in both CHA_2_DS_2_-VASc and CHA_2_DS_2_-VASc -G (below anticoagulation threshold for both): 3.9% vs. 3.7%, P=0.9, table 2, and figure 4 (method 2 in *Methods*). Indicating that the stroke risk between those down-classified and those at shared low risk was similar. These results were comparable to that observed when the reclassification table was calculated using the 4% risk threshold (eTable 7). These results were almost identical when participants that were prescribed warfarin were removed (sensitivity analysis 3, eTable 8), and similar when we recalculated this table without subtracting a point from participants who were in the bottom 25% PRS risk (sensitivity analysis 4, eTable 9) (although in this situation, as expected, no participants were down-classified).

**Figure 4:**
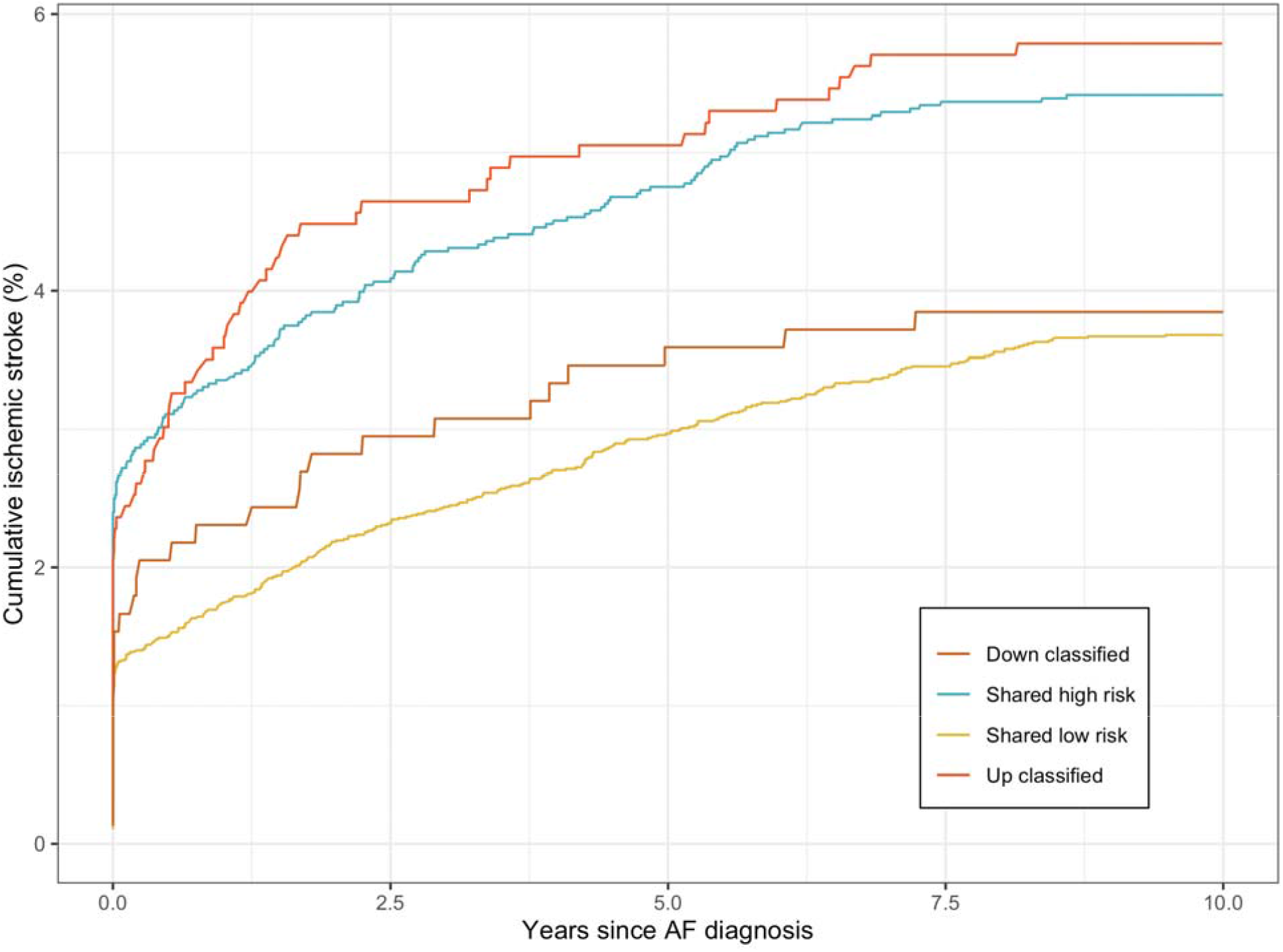
Cumulative ischemic strokes over time since AF diagnosis. Participants were grouped into four groups: Up-classified: Participants who were *below* the anticoagulation threshold using CHA2DS2-VASc, but *above* the anticoagulation threshold using CHA2DS2-VASc -G (clinical risk factors and PRS combined). Shared high risk: Participants who were *above* the anticoagulation threshold using both CHA2DS2-VASc and CHA2DS2-VASc -G. Shared low risk: Participants who were *below* the anticoagulation threshold using both CHA2DS2-VASc and CHA2DS2-VASc -G. Down classified: Participants who were above the anticoagulation threshold using both CHA2DS2-VASc, but below using CHA2DS2-VASc -G.

## Discussion

We constructed a polygenic risk score (PRS) for predicting ischemic stroke in patients with an established diagnosis of AF. Our analysis was based on over 15,000 participants in a well-conducted, prospective national biobank (UK Biobank). We extracted SNVs from the largest genome-wide association study (GWAS) (18) and combined over half a million SNVs to construct our PRS. Additionally, we built an integrated genomic and clinical risk tool, integrating our PRS with the current gold standard risk tool (CHA_2_DS_2_-VASc).

Our results show that a PRS is individually predictive of ischemic stroke in patients with an established diagnosis of AF, and this predicted risk appears independent of established clinical risk factors. The combined PRS and clinical risk tool shows significantly improved risk prediction over the current gold standard risk tool (CHA_2_DS_2_-VASc). These improvements, when applied to the large number of people with AF, translate to improved risk classification in thousands of people in the US. Nevertheless, the prediction of ischemic stroke remains challenging.

Polygenic risk scores have been produced for many cardiovascular diseases (10, 11, 14, 29, 30). A large number of PRS have been produced for coronary artery disease (CAD) (11, 14, 29) and also for ischemic stroke (10). All of these have been constructed for the population at large, compared to our PRS, which predicts an outcome (ischemic stroke) within a group of people with an established diagnosis (of AF). A minority of these previous papers have compared PRS to clinical risk factors, and even fewer have integrated PRS with clinical risk factors (and then compared this integrated tool with clinical risk factors). Abraham et al (10) constructed a PRS for ischemic stroke for all people (not just those with AF) and found a HR of 1.26, 95% CI 1.22– 1.31) and C-statistic of 0.58 (95% CI 0.57–0.59). The inclusion of all participants may explain the slightly higher HR observed by Abraham et al. Aside from their inclusion of all participants, our study differs as we compared (and adjusted for) clinical risk factors specific to stroke in AF (e.g. CHA_2_DS_2_-VASc), we integrated our PRS with CHA_2_DS_2_-VASc and determined reclassification between models.

### Implications for patients and clinicians

Our results have implications for patients, researchers and policy makers (33, 34). First, the integration of any new innovation in clinical medicine should center around patients. The integrated PRS and CHA_2_DS_2_-VASc tool significantly improves risk classification of patients; this means the tool can help identify which AF patients that will likely benefit from prophylactic anticoagulation and also potentially prevent people from unnecessary anticoagulation prescription. This latter point spares people from an increased bleeding risk, as well as the costs associated with prescription (for an individual) and the costs associated with bleeding complications (for hospital and society). To give some insight into the number of people that would be spared unnecessary bleeding risk by using the integrated PRS and CHA_2_DS_2_-VASc tool, we have extrapolated our results to AF estimates in the US. There are an estimated 5.1 million people with AF in the US (35, 36). Using the improved, integrated PRS and CHA_2_DS_2_-VASc tool, 117,000 patients with AF would have improved risk stratification. More specifically, of the 5.1 million people with AF in the US, ∼30% are eligible for prophylactic anticoagulation (from our results: table 2) (>1.5 million people). The cumulative incidence of bleeding complications from any anticoagulant (in those with AF) is estimated at 30% (37), which translates to ∼460,000 people cumulatively suffering from bleeding complications. Using the integrated PRS+ CHA_2_DS_2_-VASc tool, around 11,000 of these people will be reclassified and not be recommended anticoagulation, and thus not suffer from these bleeding complications. Similar to previous research (38), this shows that even modest improvements in risk prediction have implications for a large number of people.

Despite the improved accuracy and prediction metrics, any consideration of integrating the improved CHA_2_DS_2_-VASc -G should be cautious. More and more, medical decisions are made with ‘shared-decision making’ (39), where the risk and benefits of an intervention (or risk tool), are extensively explained with patients, and a tailored decision can be reached collaboratively. For instance, a patient with AF would begin a conversation regarding their risk of ischemic stroke and necessity for prophylactic anticoagulation. A healthcare professional would discuss the pros and cons of risk stratification via CHA_2_DS_2_-VASc, followed by the pros and cons of risk stratification via CHA_2_DS_2_-VASc -G. They may choose to highlight the >2% NRI, as well as the potential costs of genetic testing. It is likely that these pros and cons will be of varying value to different people, however with presentation of all the available data the patient’s values can lead, with the aid of a healthcare professional, to an informed decision.

### Implications for policy makers

Second, our results may be of value to policy makers. Risk tools are continually updated, mainly when new covariates are identified that improve model fit and prediction. This is evident through the history of the currently recommended CHA_2_DS_2_-VASc score. It was initially developed as the ‘CHADS_2_ tool’ (28), and with the emergence of evidence suggesting new covariates that improve the model, the tool was updated to CHA_2_DS_2_-VASc (6, 7). Our paper shows that the addition of PRS improves CHA_2_DS_2_-VASc, and future iterations of guidelines may benefit from considering its addition. This is particularly relevant due to the decreasing costs of genetic testing (40), genetic data is attained (and paid for) once, and there are multiple clinical uses once genetic data has been attained.

### Implications for researchers

Third, our study design may be of interest to other researchers. We chose to construct a PRS that a) could lead to actionable clinical changes and b) exists within current clinical practice. Currently, if a patient with AF has a CHA_2_DS_2_-VASc of score of ≥2 (in men) or ≥3 (in women), guidelines suggest they should be recommended anticoagulation (41). This represents an actionable decision threshold, based on an accepted threshold of risk. We modelled the addition of a new covariate (PRS) into an existing risk tool (CHA_2_DS_2_-VASc) at an accepted risk threshold (≥2 in men or ≥3 in women) to initiate an actionable outcome (prescription of prophylactic anticoagulation).

Furthermore, we explored reclassification between the current clinical risk tool (CHA_2_DS_2_-VASc) and our proposed integrated clinical and polygenic risk tool using two approaches. As stated in the methods and results, we used both a percentage risk threshold (method 1) as well as re-calculated each participant’s CHA_2_DS_2_-VASc score with the addition of their PRS (method 2). For both of these approaches, we found that of the participants that were up-classified (participants who were below the anticoagulation threshold using current risk stratification, but above the anticoagulation threshold using our integrated clinical and polygenic risk tool) the proportion who went on to have ischemic strokes was similar to participants who were above the anticoagulation threshold using both current risk tools and our integrated clinical and polygenic risk tool (table 2, and eTable 7). We similarly observed a comparable stroke rate for participants who were down-classified (participants who were above the anticoagulation threshold using current risk stratification, but below the anticoagulation threshold using our integrated clinical and polygenic risk tool) and for those participants who were below the anticoagulation threshold using both current risk tools and our integrated clinical and polygenic risk tool (table 2 and eTable 7). This is reassuring as it suggests that our up-classification captures those at a similar risk to those at shared high risk (and vice versa for down-classified). Lastly, we plan to make our PRS available upon publication at http://www.pgscatalog.org/.

### Study limitations

Our study should be interpreted with an understanding of its limitations. Our study was limited by the demographics of the UK Biobank. Most notably, the UK Biobank is of primarily European ancestry and we only included those of European ancestry in this study, the UK Biobank recruited participants aged 40-69 years old, and who are healthier and more affluent than the general UK population (31). Most studies that used the UK Biobank were only able to follow participants for seven years; we were able to overcome this limitation as our outcome of interest (ischemic stroke) is algorithmically defined centrally by the UK Biobank using an amalgamation of non-UK Biobank data (e.g. electronic health records from primary care etc). However, primary care data has only been released from around half of the UK Biobank participants (∼230,000 participants).

Further, it is plausible that participants were prescribed an anticoagulant after recruitment. Anticoagulant prescription could affect our study results by lowering the risk of ischemic stroke, however we believe our results remain robust for two reasons 1. Our sensitivity analyses, both adjusting for and removing participants prescribed anticoagulation at baseline, showed almost identical results to our main analysis, and this was true for both logistic and Cox regression, and 2. The prescription of an anticoagulant is likely to lead to an underestimation of the predictive power of PRS, as polygenic risk seems to mirror anticoagulation prescription (eFigure 2). This limitation is also observed in PRS for CAD (with statins) (11, 14). Furthermore, novel anticoagulants (NOACs, also known as direct anticoagulants) were not approved for stroke prophylaxis in patients with AF in the UK at the time of UK Biobank recruitment (dabigatran and rivaroxaban were approved in 2008, and apixaban in 2011) (32), nor were they used commonly in the UK until after 2015 (the first time their use was greater than warfarin) (32). Thus, NOACs were not listed in the medications code (https://biobank.ctsu.ox.ac.uk/crystal/coding.cgi?id=4&nl=1) and no participants were on NOACs at recruitment. Given NOACs have been shown to be more effective at preventing strokes in patients with AF (5), their absence from our study may actually be advantageous; we were able to capture more closely the non-intervention incidence of stroke, without the mitigating effects of NOACs. A further limitation is the inclusion of non-specific phenotype outcomes; we used ischemic stroke as our outcome rather than cardioembolic ischemic stroke (specific to AF). Unfortunately, the UK Biobank does not stratify ischemic stroke outcomes into subtypes. This may further explain why we observe slightly lower HR per 1 SD than was observed by Abraham et al (10). Similarly, stroke phenotyping presents unique challenges. Hemorrhagic and ischemic strokes can present similarly and ischemic strokes can transform into hemorrhagic strokes. It thus seems plausible that some patients are mis-phenotyped (at the UK Biobank level).

Furthermore, we used GWAS summary statistics originally derived on patients with stroke (MEGASTROKE (18)). There are no GWAS data available that are specific to patients with AF who go on to suffer from an ischemic stroke. However, we did use the cardioembolic GWAS summary statistics within MEGASTROKE. Additionally our sample size is smaller than previous PRS studies that have used the UK Biobank. This is because we only included participants with AF. A more broad weakness is the limitations of CHA_2_DS_2_-VASc; its discriminative ability to poorer than other cardiovascular risk tools (e.g. AHA/ACC’s pooled cohort equation) and hence improving on it is somewhat expected. Nevertheless, we feel integrating PRS with the current gold standard risk tool is important, even if the gold standard is sub-optimal. Lastly, a weakness of our PRS is that it was constructed from entirely common variants, and no rare variants. Although no rare, high-risk variants have been identified for ischemic stroke and the inclusion of exclusively common variants is currently common practice in PRS research, polygenic risk scores will likely become more predictive with the inclusion of rare variants.

### Future research

Our study identifies a number of research priorities. First, even with the addition of PRS, ischemic stroke remains difficult to predict. There are numerous factors that likely contribute to this. Namely, heterogeneous phenotyping and unidentified risk factors. Future research resources should focus on improving phenotyping, electronic health records, and the identification of new stroke risk factors (e.g. biomarkers, transcriptomics). Second, it would be advantageous for formal health economics studies to evaluate the cost-effectiveness of potentially implementing a combined PRS and clinical risk factors tool. Third, a tool that integrates the genetic risk of both ischemic stroke and bleeding is likely to be most useful to clinicians and patients, as they weigh up the risk and benefits of prophylactic anticoagulation. Lastly, it is vital that future GWAS, polygenic risk scores and large biobanks include non-European populations. We were unable to include non-Europeans in our study as less than 5% of UK Biobank participants are non-European. Thus the number of participants with AF that are non-European is likely to be less than 1000 and the number of participants that are non-European with AF who had an ischemic stroke is likely to be less than 15.

## Conclusion

Our PRS of over half a million SNVs is individually predictive of ischemic stroke in patients with an established diagnosis of AF, and this predicted risk appears independent of established clinical risk factors. The combined PRS and clinical risk tool (our proposed, CHA_2_DS_2_-VASc-G) shows significantly improved risk prediction over the current gold standard risk tool (CHA_2_DS_2_-VASc), however the prediction of ischemic stroke remains challenging.

### Perspectives

#### COMPETENCY IN MEDICAL KNOWLEDGE

The integration of clinical risk factors and polygenic risk score collectively had the greatest predictive accuracy to predict ischemic strokes in patients with Atrial Fibrillation.

#### TRANSLATIONAL OUTLOOK

Future studies should aim to replicate these findings in non-European ancestral populations and determine how best this integrated tool can be implemented clinically.

## Data Availability

The UK Biobank data is available upon application. We plan to publish our polygenic risk score at http://www.pgscatalog.org/ upon publication.

## Acknowledgements

A condition of the free download of the MEGASTROKE data is inclusion of the following statement: “The MEGASTROKE project received funding from sources specified at http://www.megastroke.org/acknowledgments.html“. We have also listed all MEGASTROKE authors appearing in the main author byline in our appendix, however no authors were involved in the design or conduct of this study.

## Abbreviations

CHA2DS2-VASc: Acronym of the currently recommended tool for the risk stratification of ischemic stroke in patients with AF. C = Congestive Heart Failure, H = Hypertension, A_2_ =Age (over 65 or over 75), D = Diabetes Mellitus, S = Stroke, V = Vascular Disease, S = Sex
CHA2DS2-VASc -G: A proposed term for the integrated genetic and clinical risk stratification tool, where G = Polygenic risk score.
GWAS: Genome-wide association study
AF: Atrial Fibrillation
SNV: Single nucleotide variant (polymorphism)
PRS: Polygenic risk score
SD: Standard deviation
NRI: Net reclassification index

